# Expanded statistical analysis plan for legacy and long-term effects of aspirin in the ASPREE-XT observational follow-up study of participants in the ASPREE randomized trial

**DOI:** 10.1101/2023.09.13.23295514

**Authors:** Rory Wolfe, Jonathan Broder, Andrew Chan, Anne Murray, Suzanne Orchard, Galina Polekhina, Joanne Ryan, Andrew Tonkin, Katherine Webb, Robyn Woods

## Abstract

The ASPREE randomized controlled trial (2010-2017) of 19,114 community-dwelling older adults without cardiovascular disease and significant disability compared daily 100mg aspirin to placebo. A total of 16,317 (93%) of 17,546 surviving and non-withdrawn participants agreed to continue regular study follow-up visits in the post-trial phase, named ASPREE-XT (2017-2024). We present a statistical analysis plan to underpin three main papers to report aspirin effects through to the fourth post-trial ASPREE-XT study visit with focus areas of: (1) death, dementia, and disability, (2) CVD events and bleeding, and (3) cancer. The focus of the plan is to estimate long-term (entire timespan of RCT plus post-trial) and legacy (post-trial period only) effects of aspirin in the setting of primary prevention for older individuals. Preliminary insights to these effects are presented that are based on data that has been reported to the study’s observational study monitoring board however formal data lock is not expected until October 2023.

## BACKGROUND

Between March 2010 and December 2014, the ASPREE randomized controlled trial (RCT) randomized 19,114 community-dwelling older adults without cardiovascular disease (CVD) and significant disability to daily 100mg aspirin or matching placebo (McNeil, Woods 2018). Over a median 4.7 years to the termination of intervention in June 2017, participants were followed for the occurrence of prespecified endpoint events including dementia, persistent physical disability, cancer, CVD events, major haemorrhage, depression and death. In the final 12 months of the trial, 62% of participants in the aspirin group and 64% of the placebo group were adhering to study drug (McNeil, Woods 2018 Table S6). At the study end, 29% in the aspirin group were no longer taking regular daily aspirin (study drug or open label) and 11% of the placebo group reported ongoing open label aspirin use (Smith 2023).

A total of 16,317 (93%) of 17,546 surviving and non-withdrawn participants agreed to continue in the post-trial phase, named ASPREE-XT (Ernst 2023). Their follow-up for all endpoints included annual visits and six-monthly phone calls.) Effects of long-term aspirin use on cancer and dementia are not expected to materialize for an extended period of time due to the latency periods of tumour development and preclinical cognitive decline. This has been a strong motivation for extending the follow-up of participants through ASPREE-XT.

To guide the statistical analysis of the long-term effects of aspirin, the ASPREE-XT protocol contains a statistical analysis plan which is an extension of the original trial analysis plan (Wolfe 2018). In addition, an observational study monitoring board (OSMB), appointed by the US National Institute on Aging, meets 6-monthly to oversee progress of the post-trial phase and has recommended certain analyses to be conducted. Further, in early 2023, discussion and planning commenced for three main papers to report aspirin effects through to the fourth post-trial XT study visit with focus areas of: (1) death, dementia, and disability, (2) CVD events and bleeding, and (3) cancer. This document brings together the multiple sources of input into an overarching analysis plan for these three papers. The full list of analyses outlined here is not expected to be relevant in its entirety to all three papers, and the analyses are not presented in any order of importance or preference. In addition the outlined analyses may not be exhaustive and it is anticipated that further, non pre-specified, analyses may be conducted for these main papers.

Reviews of the reporting and methods for post-trial legacy effects highlight that both direct effects (during the RCT timeframe) and legacy effects (during the post-trial period) contribute to the overall long-term effects of the intervention (Zhu et al 2021). All three types of effect may be of interest as the target of inference. It is well recognized that post-trial comparisons, even if analysed by original randomization, suffer from confounding because of selection (volunteer) bias into the post-trial follow-up.

ASPREE has already reported the direct effects of aspirin, both as a randomized comparison with placebo (McNeil, Woods 2018; McNeil, Nelson 2018; McNeil 2018) and according to actual use and non-use of aspirin (Smith 2023). The focus of this analysis plan is to estimate long-term (entire timespan of RCT plus post-trial periods) and legacy (post-trial period only) effects of aspirin in the setting of primary prevention for older individuals. Selection bias arises from: a) censoring during the trial, due to death or withdrawal, b) censoring at the end of the trial due to non-consent to participate in extended follow-up post-trial, and c) censoring during the extended follow-up, again due to death or withdrawal.

Table 1 shows contrasting results between the RCT period and the exploratory analyses from the post-trial period for certain outcomes. For example, the RCT period indicated an increased risk of cancer-related death with aspirin, whilst no risk (or benefit) was observed in the post RCT analysis. A reduction in cancer incidence and hence, shift to benefit with aspirin is hypothesized in later follow-up and there is some indication that this might be eventuating. We wouldn’t expect more than a 10-15% decrease in cancer risk with aspirin based on the findings of other trials. The unexpected elevated risk of metastatic and late stage cancer, and cancer mortality, seen in the aspirin arm in the RCT, may have abated post-trial. If confirmed, this supports the hypothesis that aspirin’s effect was a hastening of pre-exisiting, undiagnosed cancer at randomization. With longer follow-up, that early effect may give way to a decrease in incidence of cancer in the aspirin arm.

**Table 1.** Estimates of aspirin effects in RCT, unadjusted and adjusted for compliance using the rank preserving structural failure time model (RPSFTM), and unadjusted estimates of aspirin legacy effects in post-RCT period.

|  | McNeil NEJM<br>2018a,b,c |  | Smith, Am J Epi 2023 |  | OSMB report 2023 |  |
| --- | --- | --- | --- | --- | --- | --- |
| Outcome | ITT analysis using<br>the Cox PH model |  | Compliance-adjusted<br>estimates using the<br>RPSFTM |  | ITT analysis using<br>post-RCT non-<br>curated data |  |
|  | HR | 95% CI | HR | 95% CI | HR | 95% CI |
| Primary endpoint | 1.01 | (0.92, 1.11) | 1.02 | (0.84, 1.20) | 1.05 | (0.98, 1.13) |
| Death | 1.14 | (1.01, 1.29) | 1.25 | (1.00, 1.63) | 1.04 | (0.97, 1.13) |
| Dementia | 0.98 | (0.83, 1.15) | 0.93 | (0.75, 1.35) | 1.04 | (0.90, 1.21) |
| Persistent physical disability | 0.85 | (0.70, 1.03) | 0.82 | (0.64, 1.07) | 1.00 | (0.87, 1.14) |
| Major hemorrhage | 1.38 | (1.18, 1.62) | 1.74 | (1.31, 2.19) | 1.13 | (0.95, 1.36) |
| MACE | 0.89 | (0.77, 1.03) | 0.86 | (0.70, 1.08) | 1.18 | (1.02, 1.36) |
| CVD events | 0.95 | (0.83, 1.08) | 0.93 | (0.77, 1.11) | 1.13 | (0.99, 1.28) |
| Cancer | 1.04 | (0.95, 1.13) | 1.06 | (0.95, 1.19) | 0.91 | (0.82, 1.01) |
| Death from cancer | 1.31 | (1.10, 1.56) | 1.49 | (1.14, 1.91) | 0.99 | (0.85, 1.15) |

CVD – cardiovascular disease; HR - Hazard Ratio; ITT – Intention-to-treat; MACE – major adverse atherothrombotic cardiovascular events.

For MACE, a hazard ratio of 0.89 (aspirin vs placebo) was seen in the RCT, consistent with existing literature. For example, the ASCEND trial of daily low-dose aspirin in individuals with diabetes, conducted contemporaneously with ASPREE (ASCEND Study Collaborative Group 2018), showed a 12% risk reduction in serious vascular events with aspirin (with minor differences in MACE definition compared to that of ASPREE). Meta-analyses of placebo-controlled trials of low-dose aspirin both before (Antithrombotic Trialists’ Collaboration 2009), and after ASPREE, ASCEND and another contemporary trial ARRIVE (Gaziano 2018), showed similar hazard ratios for serious vascular events in those assigned aspirin (Guirguis-Blake 2022; Zheng 2019). In ASPREE, there is a notable contrast between the findings in Table 1 of MACE benefit during the ASPREE RCT (hazard ratio, HR 0.89, 95% CI: 0.77, 1.03), and the unexpected elevated risk of MACE post-trial (HR 1.18, 95% CI: 1.02, 1.36) in individuals originally randomized to aspirin compared to placebo.

These tentative insights to aspirin’s long-term effects require careful interpretation since they are based on our analyses from data that has not been properly curated and cleaned, as the posttrial ASPREE-XT follow up is still ongoing. Careful statistical analysis of locked data will be required to produce reliable estimates of aspirin effects, both as randomized and as treated.

### Defining compliance and duration of aspirin exposure

The definition of compliance used for the RCT period in Smith (2023) was based on taking regular daily aspirin for an extended period, utilising (i) annual pill count from returned bottles of study medication, and (ii) annual visit concomitant medications questionnaire responses indicating if participants were taking open-label aspirin during that year (as distinct from study medication).

Time on aspirin was estimated for each study participant in the aspirin arm by adding the number of pills taken in a participant’s last year on medication to the number of days from randomization to the beginning of that year. A period of two years without study medication intake was deemed equivalent to permanent cessation and a participant’s last year on medication was the year prior to that period.

Time on open label aspirin was estimated for participants in both treatment arms who reported use at two consecutive calendar year,s with annual visit dates for these calendar years used as starting time on aspirin (or stopping time where relevant).

For the randomized aspirin group, participants were defined as becoming non-compliant (switching) upon a permanent cessation of study medication or, if applicable, subsequent additional cessation of a contiguous period of open-label aspirin use. For the randomized placebo group, participants were defined as becoming non-compliant when they commenced open-label aspirin according to the above definition. Any switchbacks (aspirin⟵no aspirin⟵aspirin or placebo⟵aspirin⟵no aspirin) that satisfied the above definitions were ignored (fewer than 2% of participants who switched according to these definitions had a subsequent switchback).

These compliance definitions have not been extended into the XT period due to absence of study drug in that period and the addition of specific aspirin use questions in the XT data collection. A final consideration for the RCT period is an additional definition of compliance with study protocol for the entirety of the RCT (with regard to use of study drug) which is defined simply as requiring an average pill count ≥ 85% and no report of open-label aspirin, across all RCT period visits.

The following outlines the definitions for ongoing regular aspirin use that is consistent across both the RCT timeframe and ASPREE-XT.

### efinition for year by year indicators of aspirin use

Aspirin use is defined, for a given year, as either study drug use ≥ 85% (for participants in the randomized aspirin arm only), or open-label use determined by self-report ≥ 6 days/week use (specific exposure times deterimied during ASPREE-XT onwards), or concomitant medication reported aspirin (obtained during the trial and post-trial periods), for that year.

### Definition for duration of regular aspirin exposure

There may also be a need for a definition of “duration of regular daily aspirin exposure” to be used in landmark analyses of legacy effects. This would be a one-number summary of a participant’s exposure within the RCT timeframe. Hence, using year by year indicators of aspirin use, a sum of aspirin use variables would be created for visits that occurred within the RCT period. Specifically, duration in years is defined as: (i) Aspirin arm, sum of years with study medication compliance ≥ 85% and/or reported open-label aspirin use, (ii) Placebo arm, sum of years with reported open-label aspirin use. In analyses, this could be categorized, e.g., as “none”, “<2y”, “2 to <4y”, “4y+”.

These definitions are acknowledged to have limitations. Intervals between annual visits are 12 months on average but sometimes vary slightly in duration. There is a lack of information on dose for self-report and concomitant medication reporting of aspirin use (we assume low-dose) and an absence of information on frequency of use for concomitant medication reporting (although this must be ≥ 1 day per week for at least one month to be recorded; we assume ≥ 6 days/week given usual patterns of aspirin prescribing and use). Reported aspirin use, either self-report or in concomitant medication reporting, is relevant respectively to current use or the most recent one-month period (to avoid recall bias) but this means that some respondents may only have had aspirin use for a portion of the year. A sensitivity analysis is proposed using a more liberal cut-off for study medication compliance of ≥ 15%, and self-report use of ≥ 2 days/week, to create definitions that can be interpreted as some use of daily low-dose aspirin (open-label or randomized; irregular or regular) for at least a portion of the year.

### ESTIMANDS

The estimand framework is now the recommended approach by the International Committee on Harmonization (E9 (R1) Addendum 2020) for specifying statistical analysis plans of clinical trials. We have defined four estimands of interest in ASPREE and the following apply in each case:

**Endpoint** is time to the first occurrence of one of the primary or secondary endpoints of the trial (either time from randomization or time from the end of study drug intervention, as specified).

**Target population** is community-dwelling adults aged 70 years and older (65 years and older among US African Americans and Latinos) free of cardiovascular disease events, dementia, significant cognitive impairment and significant physical disability, attending primary care with their family physician / general practitioner.

**Treatment condition** is low-dose daily aspirin for an intended multi-year period and compared to placebo for a matching period (a subsidiary analysis is proposed to examine the effects of longer vs shorter period based on early vs later randomization time).

**Intercurrent events and strategies to address these: (i)** the occurrence of death (for non-fatal endpoints) and certain causes of death (for endpoints that include specific causes of death) will be treated as competing risks; **(ii)** the withdrawal of consent during the RCT and/or during XT will be accommodated using inverse probability of censoring weights (IPCW) if a substantial number of individuals are affected or, if relatively small numbers of individuals are affected, such individuals will be assumed to have withdrawn at random relative to the outcome events being analysed (i.e., noninformative censoring). These considerations do not apply to analyses of all-cause mortality which is available from public record and does not rely on having consent for continued follow-up; **(iii)** the non-provision of consent for XT follow-up will be accommodated using inverse probability weights (IPW) for analyses restricted to the post-trial period and IPCW for analyses of the entire period from randomization; **(iv)** the occurrence of cessation of aspirin use (for randomized aspirin group) or commencement of aspirin use (for randomized placebo group) will be dealt with in different ways depending on the treatment effect to be estimated (see individual estimand statements below).

**Population-level summary measure:** average treatment effect unless otherwise specified.

### ESTIMAND 1 – Effects over the long-term of a multi-year daily aspirin strategy for primary prevention in older age

**Clinical context:** Decision making with patients considering aspirin for primary prevention; what would effects be of adopting an intended strategy of multi-year regular aspirin over the long-term and specifically to 10 years?

**Analyses:** An intention-to-treat approach, i.e., according to the group to which participants were originally randomized and without reference to their actual compliance with assigned treatment or open-label aspirin, either before or after the cessation date (June 12, 2017) of the trial’s intervention phase.

For all endpoints, the primary analysis will be a Cox proportional hazards regression model with randomized treatment group as the single covariate. This model will yield ITT HRs for MACE, MI/CHD, and total/ischemic/hemorrhagic stroke, etc., for the entire cumulative follow-up from randomization to XT04.

It is unlikely that the effect of treatment will be constant over the full duration of long-term analysis time. Hence, a secondary set of analyses will compare outcomes between randomized groups using restricted mean survival time, RMST, at 10 years after randomization allowing for the competing risk of death. In a sensitivity analysis, the RMST will be estimated from Royston-Parmar models that allow for time-dependent effects of treatment by incorporating an interaction of treatment with a spline function for the cumulative hazard of events (Royston 2011).

### ESTIMAND 2 – Legacy effects of a multi-year daily aspirin strategy for primary prevention in older age

**Clinical context:** Legacy effects of a planned long-term strategy of primary prevention regular aspirin use for individuals who will survive event-free to the end of the intended multi-year strategy.

**Analyses:** By randomized group, i.e., ITT, a landmark survival analysis will be undertaken with resetting of time zero to the day after June 12, 2017 in Cox proportional hazards regression and including participants who at that landmark time, are still at risk of experiencing a first occurrence of the event of interest.

Factors possibly influencing non-inclusion in post-trial follow up will be examined by comparing descriptions of individuals included in these analyses (by compliance status at end of RCT) and individuals not included (by reason for non-inclusion: event of interest, death without event of interest, non-provision of consent for XT follow-up without event of interest). IPW for non-provision of consent will be constructed (including time in the RCT as a covariate) and adjusted for in the Cox PH models to allow for possible non-randomized differences between groups at the landmark time.

### ESTIMAND 3 – Legacy effects of adherence to a multi-year daily aspirin strategy for primary prevention in older age

**Clinical context:** Legacy effects (i.e., post-trial effects) in individuals who would comply with a long-term strategy of primary prevention regular aspirin use; for individuals who will survive event-free to the end of the intended multi-year strategy.

**Analyses:** By compliance status at the end of the RCT, i.e., as-treated, a landmark survival analysis will be undertaken with resetting of time zero to the start of the day after 12 June 2017 (events occurring on that day will be attributed time 0.5 days for analysis). Cox proportional hazards regression will be applied including participants who at the landmark time are still at risk of experiencing a first occurrence of the event of interest.

Factors that influenced selection out of post-trial follow up for reasons other than death (considering only selection prior to commencement of post-trial follow-up, and including the reason of non-provision of consent to participate in the XT study) will be adjusted for by inverse probability weights of non-inclusion in XT to weight participants in XT to better represent the survivors from the originally randomized cohort.

The use of alternative as-treated approaches, specifically, complier-average causal effects, may also be considered. Thus, legacy effects will be estimated for individuals having the propensity to comply fully with a multi-year regular aspirin primary prevention strategy.

### ESTIMAND 4 – The effects of ongoing use of daily aspirin for primary prevention of CVD in older age

**Clinical context:** Decision making with patients aged >70y in general practice setting considering aspirin for primary prevention; what would be the effects of adopting and adhering to multi-year regular aspirin use over the long-term.

**Analyses:** Approaches such as excluding participants who “switch” to become non-adherent with intended intervention, or censoring them at the time of switching are problematic because switchers are likely to be prognostically different from non-switchers (Latimer 2016).

Suitable methods to enable as-treated analyses while allowing for factors influencing switching are: marginal structural models (MSM) using inverse probability of censoring weighting (IPCW) for treatment switches; and (ii) rank preserving structural failure time models (RPSFTM) using g-estimation.

The precise times of treatment switching are unknown, and the effect of taking aspirin will be estimated using a RPSFTM applied to data from randomization through XT04 with estimates of cumulative time “on” aspirin use and cumulative time “off” (whether through adherence to randomized study drug or use of open label aspirin during the RCT or afterwards). A similar simplifying approach to estimating time on treatment (up to switch time) could be considered for use with the MSM approach.

### HYPOTHESES

A number of specific hypotheses have been proposed to examine possible influences on estimates of aspirin-related treatment effects. The hypotheses pre-specified here are not intended to be exhaustive in this regard. We anticipate there will be important post-hoc hypotheses and further exploratory analysis to fully understand the insights to aspirin effects that are obtained from the analyses to be conducted under this analysis plan.

### HYPOTHESIS A: Aspirin cessation causes some (short-term) unmasking of cardiovascular harm

If we were truly testing aspirin withdrawal in an RCT, we would take those initially randomized to aspirin, and then randomize them at the end of RCT phase to aspirin withdrawal or not.

**Analyses:** Emulate a deprescribing trial of aspirin by constructing a comparison of those continuing aspirin use to those ceasing aspirin use at the end of the RCT period, adjusted for propensity of continuing/ceasing. Look at outcomes in the short-term period thereafter (6-12 months: returning to normal platelet aggregation may cause clotting on established unstable lesions) and augment to also examine longer term to XT04. To avoid possible confounding due to higher CVD risk for those who continue aspirin use, remove individuals who experienced any CVD type events (angina, mini-stroke etc) during the trial noting the limitation that these clinically less severe events were captured only by self-report or medical record review.

### HYPOTHESIS B: High-risk participants randomized to placebo would have had their event during ASPREE, whereas in similar high-risk participants randomized to aspirin, such events would have been prevented (or delayed until the post-trial period)

**Analyses:** Examine participant characteristics (at the ASPREE-XT baseline for those who continued follow-up) for those who ended up having a CVD event/MACE during the XT period, compared to those who did not, and by whether they were originally randomized to aspirin or placebo. Subgroup analyses by baseline CVD risk will be undertaken.

### HYPOTHESIS C: Men will reach most outcomes at different rates and overall frequency than women

The risk-benefit profile of aspirin may be differentially affected by sex due to factors such as lower rates of hemorrhage in women and greater risks of stroke and myocardial infarction in men.

**Analyses:** Subgroup analyses by sex will be undertaken. See below for a full listing of the protocol-specified subgroups and proposed additional subgroups.

### HYPOTHESIS D: Anti-cancer effects of aspirin require a certain duration of exposure in order to occur

It has been found that anti-cancer effects of aspirin require only 3-5 years of treatment before observing a later legacy benefit with longer follow-up (Rothwell 2012).

**Analyses:** Subgroup analysis by duration of potential aspirin treatment in the trial phase. Overall and also further subdivided by duration of regular aspirin use prior to the trial.

### HYPOTHESIS E: Aspirin’s effect on cancer is to hasten cancer in people who already have (undetected) tumour growth when they start on aspirin, and that with longer follow-up, cancer incidence is reduced within the aspirin arm

**Analyses:** Analyses will be conducted to exclude cases of cancer that were diagnosed within one year of randomization, and examine long-term effects according to both randomized arm and compliance status.

## ADDITIONAL ANALYSES

Describe aspirin use and additional medications including anti-thrombotics, before and after the intervention phase (overall and by preceding CVD event). For example, aspirin use post RCT has been observed to be higher in participants in the US than Australia (as it was pre-trial), and has been reducing in prevalence the longer after the trial our follow-up continues. Post-trial aspirin use is similar between randomized aspirin and placebo groups (approximately 15% in each). Post-trial aspirin use and anticoagulant use is much higher in participants who have had a preceding CVD event.

For disability-free survival and all-cause mortality, Kaplan-Meier estimates of cumulative incidence will be plotted for the two randomized groups. For all other outcomes, estimates of cumulative incidence (Aalen-Johansen cause-specific cumulative incidence function) will be plotted for the two groups having been obtained from group-stratified models in which death (from causes other than any causes incorporated in the outcome of interest) will be considered a competing risk.

Secondary analyses may include adjustment for baseline (time of randomization) characteristics that are thought to be strongly prognostic of outcome in this population (e.g., age, sex, pre-trial prior regular aspirin use and comorbidities including cognitive function).

Additionally, for the endpoints for which it is envisaged that aspirin may have an early effect that reduces in magntiude during ASPREE-XT (specifically major hemorrhage and CVD/MACE), a generalized (“combined”) test of treatment effect, comparing RMST difference at 10 equally spaced intervals of time between 3 years post-randomization (approximately the minimum duration of scheduled follow-up in ASPREE) and the time of the latest occurring event in ASPREE-XT (anticipated to be at approximately 11-12 years) will be used to test for the existence of a treatment effect at some point during that time interval while controlling for the multiplicity of the time points being employed (Royston 2016).

### Subgroup analyses

Analyses will also be undertaken within subgroups to examine for evidence of variation in the treatment effect. These subgroup analyses will be based on appropriate interaction terms in the relevant regression model for each outcome of interest (Cox proportional hazards regression model, Royston-Parmar parametric model, or landmark model). The p-values for these interaction terms will be used to test for heterogeneity of treatment effect of aspirin between subgroups. There will not be any multiplicity adjustment when calculating p-values for subgroup analyses. Subgroup analyses will be labelled as exploratory in the reporting of longitudinal outcomes based on previous ASPREE treatment groups, and effect estimates and confidence intervals for each subgroup will be reported along with the p-value for the test of interaction. The subgroups are defined using baseline (time of randomization) information, divided into pre-specified during ASPREE and new subgroups, and include:

### Subgroups specified in ASPREE-XT protocol

**a) Sex: Males versus females**

**b) Age:** (i) >/< study median (74 years) and (ii) categories 65-69, 70-74, 75-79, 80+y

**c) Country:** US versus Australia

**d) Ethnicity:** Whites in Australia, Whites in US, African Americans, Hispanics, Other (where ‘other’ will include Native Americans, Asians and Aboriginal Australians)

**e) Diabetes:** Presence/absence of diabetes as self-report, elevated fasting blood glucose [≥ 126 mg/dL (US) or ≥ 7mmol/L (Australia)] or prescribed medication for diabetes

**f) Hypertension:** Hypertensives versus non-HT. HT defined as those who are on treatment for high blood pressure or those with blood pressure recorded above 140/90 mmHg.

**g) Smoking:** Current versus Never or Former smokers

**h) Prior aspirin use**: self-reported regular use of aspirin immediately prior to baseline

**i) Frailty**: Frail (3+) versus Pre-frail (1-2) versus Not frail (0). Number of criteria satisfied, from Fried criteria for frailty.

**j) Personal history of cancer**: Any history of cancer at baseline other than non-melanoma skin cancer

**k) Hypercholesterolemia (dyslipidemia)**: self-reported use of statin at baseline or elevated cholesterol (either serum TC ≥ 240 mg/dL [≥ 6.2 mmol/L for participants in the US] or ≥ 212 mg/dL [≥ 5.5 mmol/L; for participants in Australia] or LDL >160 mg/dL [>4.1 mmol/L])

**I) BMI**: WHO criteria (kg/m^2^): underweight (<20), normal (20-24), overweight (25-29), obese (≥ 30)

**m) CKD**: eGFR <60 ml/min/1.73m^2^ or uACR ≥ 3 mg/mmol or renal transplant or routine dialysis.

**n) Waist circumference**: WHO criteria, >88 cm for women and >102 cm for men.

**o) Polypharmacy**: taking 5 or more medications.

**Additional subgroups:**

**p) Categories of fatal and non-fatal CVD risk** using the SCORE2-OP equation (SCORE2-OP working group and ESC Cardiovascular risk collaboration 2021) calculated (i) at randomization, (ii) recalculated at XT01 for legacy effect estimation.

**q) Randomization after 12 June 2013** (i.e., <4y of aspirin study drug possible) compared to randomization on or prior to 12 June 2013 (i.e., ≥ 4y of aspirin study drug possible, hypothesized to maximize legacy effects).

**r) Cross-classification of high blood pressure and use of anti-hypertensives at baseline:** refer to subgroup f.

**s) Cross-classification of elevated cholesterol and use of statins at baseline:** refer to subgroup k.

**t) Years of education:** <12, 12-15, 16+.

**u) Disability-free survival prediction categories** using Neumann-Thao-Murray equation (Neumann, Thao, Murray 2022) calculated (i) at randomization, (ii) recalculated at XT01 for legacy effect estimation.

**v) Frailty Index**: as defined in Ryan (2022); see also subgroup i.

### Further subgroups likely to be the subject of separate manuscripts

**Depression**

**LPA genetic status**

**CHIP status**

**APOe status**

**GDF15 value**

**Other inflammatory biomarkers, dementia plasma biomarkers, etc**.

## Data Availability

All data produced in the present study are available upon reasonable request to the authors

https://ams.aspree.org/public/

## ACKNOWLEDGEMENTS

We thank a wide group of ASPREE investigators for many discussions that have preceded and informed this analysis plan. This group includes but is not limited to: Mike Ernst, Sara Espinoza, Brenda Kirpach, Raj Shah, Jeff Williamson, Lawrie Beilin, John McNeil, Mark Nelson, Chris Reid, Nigel Stocks.

We are grateful to Dr Tom Sullivan of the South Australian Health & Medical Research Institute, Dr Ian Marschner of the NHMRC Clinical Trials Centre at University of Sydney and Dr Thao Le Thi Phung of Monash University for helpful comments that led us to improve the final version of the plan.

ASPREE was funded by grants (U01AG029824 and U19AG062682) from the National Institute on Aging and the National Cancer Institute at the National Institutes of Health, by grants (334047 and 1127060) from the National Health and Medical Research Council of Australia, and by Monash University and the Victorian Cancer Agency. We are also grateful to the ongoing participation of the individuals who make up the ASPREE cohort and their families, general practioners and primary care physicians.

